# SARS-CoV-2 Viral RNA Load Dynamics in the Nasopharynx of Infected Children

**DOI:** 10.1101/2020.08.31.20185488

**Authors:** Kai-qian Kam, Koh Cheng Thoon, Matthias Maiwald, Chia Yin Chong, Han Yang Soong, Liat Hui Loo, Natalie Woon Hui Tan, Jiahui Li, Karen Donceras Nadua, Chee Fu Yung

## Abstract

It is important to understand the temporal trend of pediatric *severe acute respiratory syndrome coronavirus 2* (SARS-CoV-2) viral load to estimate the transmission potential of children in schools and communities. We determined differences in SARS-CoV-2 viral load dynamics between nasopharyngeal samples of infected asymptomatic and symptomatic children. The daily cycle threshold values of SARS-CoV-2 in the nasopharynx of a cohort of infected children were collected for analysis. Among 17 infected children, 10 (58.8%) were symptomatic. Symptomatic children, when compared to asymptomatic children, had higher viral load (mean cycle threshold on day 7 of illness 28.6 versus 36.7, p = 0.02). Peak SARS-CoV-2 viral loads occured around days 2-3 of illness/days of diagnosis in infected children. After adjusting for the estimated date of infection, the higher SARS-CoV-2 viral loads in symptomatic children remained. We postulate that symptomatic SARS-CoV-2-infected children may have higher transmissibility than asymptomatic children. As peak viral load in infected children occurred in the early stage of illness, viral shedding and transmission in the pre-symptomatic phase probable. Our study highlights the importance of screening for SARS-CoV-2 in children with epidemiological risk factors, even when they are asymptomatic in order to improve containment of the virus in the community, including educational settings.

**Key points:**

- Symptomatic children had higher SARS-CoV-2 viral loads in the nasopharynx than asymptomatic children, which may indicate that symptomatic children have higher transmissibility.
- Peak SARS-CoV-2 viral loads occurred early around 2-3 days post symptom onset iin children and therefore, pre-symptomatic transmission of the virus is probable.
- Symptom based screening for SARS-CoV-2 may not be effective in diagnosing coronavirus disease 2019 (COVID-19) in children as a proportion of children may be asymptomatic or pauci-symptomatic.
- Children with high epidemiological risk factors should be screened or isolated as they may be carriers of the virus and contribute to transmission.

---

The viral load of the novel coronavirus, *severe acute respiratory syndrome coronavirus 2* (SARS-CoV-2), has been reported to peak within the first week of disease in throat and sputum samples in the adult population [1-2]. The viral loads in a small group of asymptomatic infected adults were also shown to be similar to those of symptomatic infected adults, implying a transmission potential of asymptomatic patients [2]. A proportion of COVID-19-infected children are known to asymptomatic or have mild to moderate disease [3-5]. Previously, we reported a paucisymptomatic SARS-CoV-2-infected infant who presented with high viral load prior to symptom onset, viral shedding up to day 18 of illness and significant environmental viral contamination [6, 7]. A recent study in Korea also suggests that SARS-CoV-2 can be detected in children for a mean of more than 2 weeks and a proportion of the infected children are asymptomatic despite high viral load [8]. There is a need to improve the understanding of the viral load dynamics of SARS-CoV-2 in infected children so as to postulate the role of children in the transmission of COVID-19 in schools and the community. In this study, we analyzed the daily trends of SARS-CoV-2 viral load from nasopharyngeal samples of infected symptomatic and asymptomatic pediatric patients.

KK Women’s and Children’s Hospital (KKH) is an 830-bed hospital that provides care for approximately 500 children’s emergency daily attendances and 12,000 deliveries per year. It is the primary hospital for evaluation and isolation of COVID-19 in the pediatric population in Singapore. A line list of confirmed pediatric cases who presented to our institution from 23 March 2020 to 5 April 2020 was extracted from hospital records. Confirmed cases that were diagnosed by positive SARS-CoV-2 PCR from nasopharyngeal swabs using the real-time reverse transcription polymerase chain reaction (rRT-PCR) for the E gene were included. Nasopharyngeal swabs were taken daily from the confirmed cases. A cycle threshold (Ct) of 45 is considered to be undetectable for the virus. Age, gender and the Ct values of all nasopharyngeal swabs for SARS-CoV-2 for each child were also obtained. Most of these children were contacts of confirmed cases in their household. In our institution, children were considered to have recovered from COVID-19 and discharged from the hospital when they had negative SARS-CoV-2 PCR results from 2 nasopharyngeal swabs on consecutive days.

The Ct values were reported in relation to the day of illness or day of diagnosis for symptomatic and asymptomatic patients, respectively (Figure 1A). As an additional analysis to mitigate bias in timing of detection between symptomatic and asymptomatic cases, we incorporated an estimated day of infection in Figure 1B. The children were most likely to have been infected by their household confirmed cases during this early period of the pandemic in Singapore with detailed contact tracing and testing. Therefore, we used the date of symptom onset for index household COVID-19 case as the estimated day of infection for this additional analysis. If this information was missing, we used the average duration calculated from patients with the information available.

**Figure 1:**
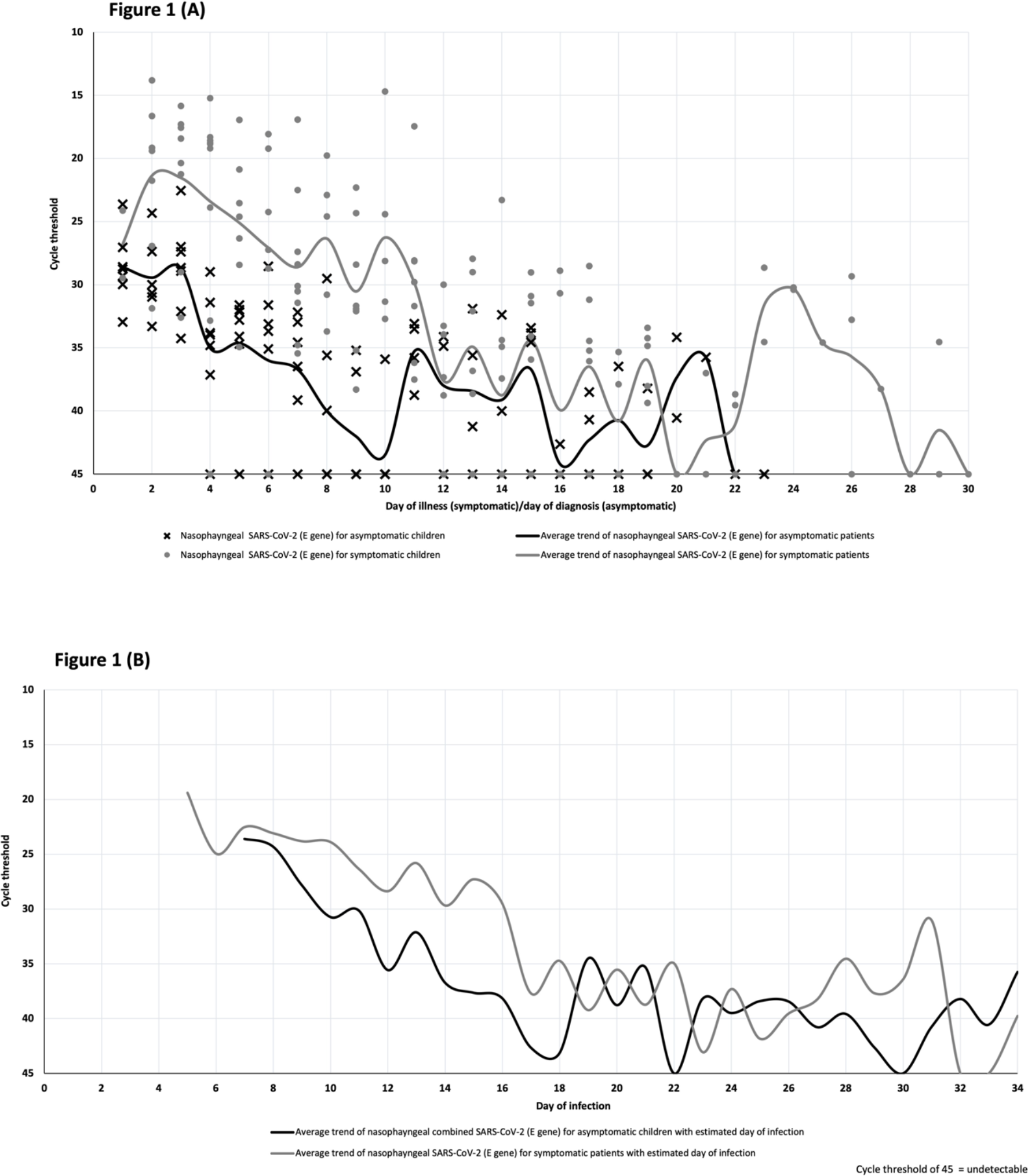
SARS-CoV-2 temporal viral load dynamics in the nasopharynx of pediatric COVID-19 patients. (A) Plotted against day of illness (symptomatic children) or day of diagnosis (asymptomatic children). (B) Plotted against estimated day of infection. SARS-CoV-2: *severe acute respiratory syndrome coronavirus 2*

Nasopharyngeal swabs were collected using Mini UTM Kits (Copan, Brescia, Italy) with flocked swabs and 1 mL of universal transport medium. From this medium, 200 μL was used for extraction of viral nucleic acids using the EZ1 Virus Mini Kit v2.0 (Qiagen, Hilden, Germany) into 60 μL of eluate. rRT-PCR targeting the SARS-CoV-2 E gene was performed according to the method by Corman et al [9]. All reactions were run on a QuantStudio 5 instrument (ThermoFisher Applied Biosystems, Foster City, CA, USA). A volume of 5 μL was used in PCRs, and with conversion factors, this represented 1/60th of the swab contents per reaction. The positive control consisted of a plasmid with a SARS-CoV-2 E gene insert, adjusted to 1000 copies per reaction. During the study period, the mean Ct value of all positive control PCRs was 29.86 and the standard deviation was ± 0.68. Thus, with the conversion factor, a Ct value of 29.86 corresponded to approximately 6 x 10^4^ virus genome copies per swab. All first-time positive results for each individual patient were confirmed by a second PCR assay, the Fortitude PCR kit (A*Star, Singapore) on a CFX96 thermocycler (Bio-Rad, Hercules, CA, USA).

Continuous variables were normal in distribution; they were expressed as mean (range) and compared with 2 samples T test. Categorical variables were expressed as numbers (%). A two-sided α of less than 0.05 was considered statistically significant. Statistical analyses were done using the SPSS software, version 23 (IBM, Armonk, NY, USA). The average Ct values for each day of illness and diagnosis were obtained for the asymptomatic and symptomatic children. The graphs were drawn to compare the average Ct trends on each day of illness/diagnosis between the two groups. The study was approved by the institutional ethics review board. Written informed consent was waived in light of the need to inform public health outbreak control policies.

From 23 March to 5 April 2020, 17 children with confirmed COVID-19 via rRT-PCR E gene assay were cared for as inpatients at our institution. The median age of the cohort was 7.7 years old (range: 0.3 to 15.8 years old). In our cohort, 10 (58.8%) of the children were symptomatic. All symptomatic children had a mild illness with upper respiratory tract infection and their symptoms resolved by day 5 of illness. No complications or evidence of pneumonia were observed. All asymptomatic children remained well with no development of symptoms until discharge.

The mean duration of viral shedding was about 16 days (range: 3-29 days) in our cohort. Symptomatic children had longer durations of viral shedding but this was not statistically significant (mean 17 days versus 14 days, p = 0.48). Higher viral loads were observed in symptomatic children when compared to asymptomatic children (mean cycle threshold on day 7 of illness 28.6 versus 36.7, 95% CI = 1.9 to 14.3, p = 0.02). Peak SARS-CoV-2 viral loads occured around days 2-3 of illness in symptomatic children or days of diagnosis in asymptomatic children, respectively (Figure 1A). After adjusting for the estimated date of infection, SARS-CoV-2 viral loads remained higher in symptomatic children when compared to asymptomatic children, but the durations of viral shedding between the two groups were similar (Figure 1B).

We present the daily nasopharyngeal SARS-CoV-2 Ct values of the asymptomatic and symptomatic infected children in our cohort. The mean duration of viral shedding was 16 days and the longest duration of viral shedding was 30 days in a previously symptomatic child. The detection of early viral peaks at days 2-3 of illness in our pediatric cases concurs with data from limited paediatric studies [10, 11]. This raises the possibility that children with COVID-19 may transmit the virus to others during the early stage of the illness. Although we were unable to establish if the virus was already detectable in high viral loads in the pre-symptomatic phase in our cohort of patients, the high viral load in our symptomatic children in the very early stage of illness implies the transmission potential of pre-symptomatic children if they are not identified and isolated in the pre-symptomatic phase.

Contrary to reports of COVID-19-infected adults showing similar viral load trends in asymptomatic and symptomatic infected individuals [2], our findings revealed that symptomatic COVID-19-infected children may have higher viral loads in the initial stage of illness than asymptomatic children. Mi et al recently reported similar findings from a smaller cohort in South Korea but daily swab data was not available [11]. Importantly, even after we accounted for the estimated day of infection for both symptomatic and asymptomatic children, the viral loads continued to be higher in symptomatic children. This suggests that symptomatic children may have a higher risk of transmitting the virus than asymptomatic children. This finding and our daily viral load data could be used to guide modelling work to help undertand the role of children in driving transmisison in schools and the wider population, in order to inform public health interventions including future vaccination policies.

We found that the majority of patients had detectable virus even on day 7 of illness/diagnosis and the mean duration of viral shedding was 16 days. If testing resources need to be preserved, we suggest that infected children do not need to be tested for the virus for at least the first 7 days of illness/diagnosis to confirm they no longer need to be isolated unless clinically indicated. Isolation of pediatric COVID-19 cases to prevent ongoing transmission may need to continue for at least 2 to 3 weeks. Children with epidemiological links to confirmed cases in the household and schools should be placed under quanratine for a least 2 weeks, as these children may shed the virus during the asymptomatic or pre-symptomatic phase.

Although our sample size is limited, daily nasopharyngeal specimens were taken for every case which accounted for individual level variations in viral shedding patterns in infected children. The findings from our cohort may not be representative of other pediatric populations but our cohort included multi-ethnic patients (Chinese, Malay, Indian, Eurasian ethnicities). Our PCR assay was not set up to be exactly quantitative, but approximate viral loads can be gleaned from Ct values, using the dilution factors and the fact that positive controls contained 1000 copies of a plasmid per reaction. As per all observational studies, it is difficult to ascertain the exact day of acquisition for the infected children. For our secondary analysis (Figure 1B), we assumed that the children were infected on the onset date of the index household COVID-19 case. Epidemiological data from Singapore’s systematic contact tracing, testing and isolation policy supported this assumption during the early phase of the pandemic [12].

In conclusion, our study found that symptomatic infected children have higher viral RNA loads than asymptomatic children. We also detected peak viral load occurred very early in children within about 2 to 3 days of infection which would suggest probable potential for transmission before symptom onset. These findings highlights the importance of screening for SARS-CoV-2 in children with epidemiological risk factors, even when they are asymptomatic in order to improve containment of the virus in the community including educational settings.

## Data Availability

The readers can contact the corresponding author if they want access to the resources and data necessary to replicate the findings discussed in this paper.

## Acknowledgement

The authors would like to thank all the staff of the Microbiology Section, Department of Pathology and Laboratory Medicine, KK Women’s and Children’s Hospital for their dedication and commitment in the challenging working conditions in this COVID-19 pandemic.

